# Nocturnal Physiological Associations with Agitation Occurrence and Severity in Dementia: An Explanatory Study Using Contactless Sleep Sensing

**DOI:** 10.64898/2026.02.27.26346707

**Authors:** Zhen Liu, Marta Bono, Ajda Flisar, Robbe Decloedt, Maarten Van Den Bossche, Maarten De Vos

## Abstract

**Background:** Agitation is a common and burdensome neuropsychiatric symptom in dementia that fluctuates from day to day, but objective tools for short-term risk stratification are limited.

**Objective:** We aimed to investigate whether nocturnal physiological signals from unobtrusive under-mattress sensors predict next-day daytime agitation and whether associations differ for agitation occurrence versus severity.

**Methods:** We extracted cardiorespiratory, movement, and sleep-proxy features from two long-term care cohorts (*N*=55; 333 nights) and one external home-monitoring cohort (*N*=17; 801 nights). A two-part mixed-effects framework was used to model next-day agitation episodes.

**Results:** Lower nocturnal respiratory rate and greater activity variability were independently associated with higher odds of next-day agitation occurrence, whereas no sleep-derived feature was FDR-significant for agitation severity. Exploratory subtype-specific analyses yielded more FDR-significant associations for motor agitation than for verbal agitation. Respiration-related associations were also observed in the external cohort.

**Conclusions:** Passive sleep monitoring identified physiologically interpretable sleep markers associated with next-day agitation occurrence, supporting prospective evaluation of under-mattress sensing as a source of short-term risk information for dementia care.

## 1. Introduction

Agitation is among the most frequent and clinically challenging behavioral and psychological symptoms of dementia (BPSD) [1]. It occurs across dementia diagnoses and commonly includes excessive or inappropriate motor activity, verbal agitation, aggression, and resistance to care [2, 3]. Beyond its immediate impact on people living with dementia (PLWD), agitation is associated with caregiver burden, safety incidents, increased healthcare use, and earlier institutionalization, making it a key target for monitoring and management in dementia care [4, 5].

Pharmacological interventions for agitation, including antipsychotics and sedatives, are associated with clinically important risks in PLWD, including adverse events and increased mortality [6]. Accordingly, current management emphasizes identifying triggers, addressing unmet needs, and prioritizing non-pharmacological approaches. However, implementation in real-world care settings remains difficult because of limited staffing, incomplete contextual information, and the episodic, fluctuating nature of symptoms [4]. Conceptual frameworks such as the Unmet Needs Model further suggest that agitation may arise from interactions between neurodegenerative vulnerability and modifiable precipitating factors (e.g., discomfort, pain, environmental stressors), highlighting the importance of identifying actionable antecedents in daily life [7].

Sleep is a promising and potentially modifiable upstream factor in this context. Sleep disturbances are highly prevalent in Alzheimer’s disease (AD) and related dementias across both community and long-term care settings [8–10]. Beyond being a common comorbidity, sleep disruption may interact bidirectionally with AD pathophysiology and downstream neurobehavioral function, providing biological plausibility that poor sleep may amplify next-day emotional distress and behavioral dysregulation [11, 12]. More broadly, experimental evidence indicates that sleep loss alters affective brain circuitry by reducing prefrontal regulatory control over limbic responses, offering a plausible framework for how disturbed sleep may heighten emotional reactivity and behavioral dysregulation [13]. Together, these observations support a day-to-day perspective in which nightly sleep features may shape subsequent daytime agitation risk.

However, establishing the sleep-agitation relationship in real-world dementia settings remains challenging because both constructs are difficult to measure. Although polysomnography (PSG) provides rich physiological information, it is resource-intensive, intrusive, and may not reflect habitual sleep patterns in naturalistic environments [14]. Actigraphy is more scalable but provides limited physiological specificity beyond movement-derived proxies [15–17]. Agitation, in turn, is often measured using retrospective caregiver reports or intermittent observational ratings, which are vulnerable to recall bias and may miss short-term symptom fluctuations [18, 19]. Recent advances in passive in-home sensing (e.g., under-mattress monitoring) provide a promising opportunity for low-burden, longitudinal characterization of nocturnal physiology in everyday dementia care contexts [20–22].

Even with these sensing tools, the sleep-agitation relationship remains insufficiently characterized. Several studies using similar sensors have focused on other outcomes, such as sleep phenotyping or long-term disease progression, rather than day-to-day agitation [22, 23]. Other work predicting agitation has relied on multimodal inputs spanning sleep, daytime activity, and environmental factors over multi-day windows, making it difficult to isolate the specific contribution of nightly sleep or capture rapid day-to-day changes [24]. A proof-of-concept study [18] showed that integrating behavioral, environmental, and sleep sensing can help characterize and predict agitation in care settings; however, the sleep component relied on a single composite score, and the small sample limited generalizability. More broadly, although sensor-based agitation detection and prediction is an active area, relatively few studies have examined sleep-derived signals in isolation, and the literature remains fragmented with respect to sensing modalities, outcome definitions, and validation strategies [25, 26]. Collectively, these gaps motivate rigorous, temporally aligned analyses that isolate the contribution of nightly sleep to next-day agitation while accounting for repeated measures and confounding.

Therefore, we aimed to quantify the association between objectively measured nocturnal sleep characteristics and next-day daytime agitation using longitudinal, real-world data and analytic methods designed for hierarchical and zero-inflated outcomes. Specifically, we evaluated whether sleep-derived physiological and behavioral features predicted (i) agitation occurrence and (ii) agitation severity among agitation-positive days, while adjusting for key confounders and accounting for between-subject heterogeneity. By focusing on a night-to-next-day window and low-burden monitoring suitable for realworld care settings, this study aimed to characterize interpretable sleep–agitation associations that may inform future risk-stratification studies and the design of prospective care-workflow evaluations. A schematic overview of the study is shown in Figure 1.

**Figure 1:**
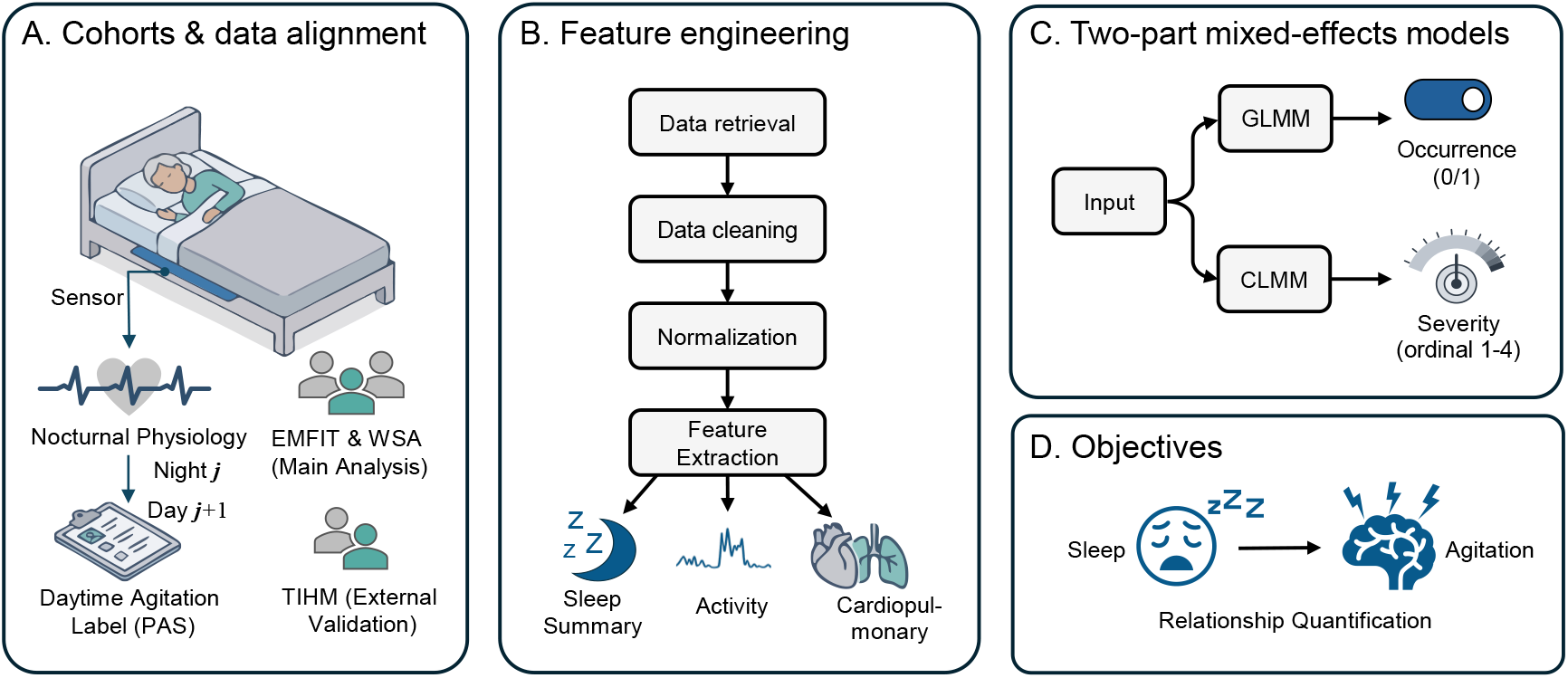
Study Design. (A) cohort structure and night-to-next-day alignment between nocturnal sensor-derived physiology (night *j*) and next-day daytime agitation labels (day *j* + 1), with EMFIT and WSA used for primary analysis and TIHM for external evaluation; (B) data processing and feature extraction; (C) two-part mixed-effects modeling of agitation occurrence (GLMM) and severity among agitation-positive days (CLMM); and (D) study objective to quantify sleep–agitation relationships.

## 2 Methods

### 2.1 Study design and data sources

This study utilized data from three distinct cohorts to ensure robustness and generalizability. The primary dataset, comprising cohorts “EMFIT” and “WSA”, was collected at “Cog K”, a specialized neuropsychiatric ward within the University Psychiatric Centre (UPC) KU Leuven [19, 27]. This unit treats patients with dementia exhibiting severe BPSD, corresponding to Tiers 6 and 7 of Brodaty’s seven-tiered care model [28]. Using convenience sampling, data were collected via unobtrusive under-mattress sensors integrated into the daily clinical workflow and retrieved via cloud APIs. Further protocol details are available in the study by Davidoff et al. [19]. The specific monitoring configurations for each cohort were as follows:

- **Cohort “EMFIT”** involved one week of continuous monitoring using the Emfit QS^®^ sensor, paired with a hybrid signal- and event-contingent ecological momentary assessment (EMA) strategy. Annotations were collected daily, averaging 6.3 labels per day (SD = 2.7).
- **Cohort “WSA”** comprised two weeks of monitoring using the Withings Sleep Analyzer^®^ (WSA), supported by a high-density event-contingent EMA protocol. This dataset exhibited a high annotation density, with a daily average of 22.1 events (SD = 10.2).
- **Cohort “TIHM”** is a public in-home remote monitoring dataset of people over 50 years old with dementia or mild cognitive impairment [29]. It was used for external validation. Eligibility required a study partner or caregiver who had known the participant for at least six months and could participate in research assessments. Individuals with unstable mental states, including severe agitation, were excluded. The released agitation label is a binary clinically validated health-event label generated after rule/model-assisted alert triage and monitoring-team validation. This label differs from the EMA-based occurrence outcome used in the WSA/EMFIT cohorts. While the original TIHM dataset includes 56 subjects monitored for an average of 50 days in a home setting with twice-daily annotations, only the subset of 17 subjects containing valid sleep data was included in this analysis.

Both devices employ an under-mattress pressure sensor and ballistocardiography (BCG) to derive heart rate (HR) and respiratory rate (RR) and to estimate in-bed movement/activity (Act) from the same signal, without requiring the user to wear a sensor or to have direct skin contact. Our exported data contains continuous HR/RR/Act time-series at high temporal resolution (WSA: 1-min; EMFIT: 4-s). Notably, although both of our proprietary datasets include activity metrics (WSA: 0–255; EM-FIT: 0–32768; both right-skewed), the public TIHM dataset does not provide accessible activity data.

Agitation was quantified using the Pittsburgh Agitation Scale (PAS) [30], which evaluates severity on a scale of 0–4 across four behavioral subtypes: motor, verbal, aggressive, and resistance to care. The PAS scores were manually recorded during the day and matched with sleep data from the preceding night. Agitation occurrence was defined as a binary variable equal to 1 when any PAS subtype score exceeded 0 during the following day. Furthermore, to capture the intensity of behavioral disturbances, agitation severity was defined as daily maximum PAS among agitation-positive days. Daily maximum PAS was the highest of the daily maximum motor, verbal, aggressive, and resistance-to-care scores, preserving the original 0–4 PAS severity scale. The TIHM dataset provided only agitation occurrence labels. Analyses of agitation severity were therefore restricted to the WSA and EMFIT cohorts.

### 2.2 Ethical considerations

Ethical approval for this study was granted by the Ethics Committee Research of University Hospitals Leuven (reference ID S62882). Given the participants’ diagnosis of dementia, written informed consent was primarily secured through legal representatives. However, to ensure patient inclusion, the study details were explained verbally in a manner commensurate with each patient’s cognitive abilities. All patient data were pseudonymized and handled with strict confidentiality. Participation was voluntary and uncompensated, with the explicit understanding that the decision to enroll would not impact the standard of care received during admission.

### 2.3 Data preprocessing

#### Time-Series

Non-contact sleep sensors have demonstrated good reliability in the long-term, continuous collection of vital-sign data from older adults and PLWD [20, 21]. The sleep-related metrics they provide, including sleep heart rate (HR) and breathing rate (RR), show good concordance with the gold standard PSG. Moreover, coarse sleep-wake classification (binary Sleep-Wake) tends to exhibit higher PSG agreement than more fine-grained four-class staging (Wake, Light, REM, Deep). Accordingly, this study computes sleep features from sleep-wake, HR, RR, and activity time series.

A median filter (three-minute window) was applied to raw HR and RR time-series data to suppress non-physiological spikes. For activity data, a two-step process was used: (i) Winsorization at the 1st and 99th percentiles within each device type to cap extreme outliers; (ii) Log-transformation (log(*x*+1)) to correct skewness. For continuous features such as HR, RR, and activity, we applied device-wise robust Z-score normalization using device-specific medians and interquartile ranges (IQRs).

#### Feature Extraction

We implemented a comprehensive feature extraction pipeline to characterize the multidimensional nature of nocturnal physiology and behavior. From the sleep-wake sequence, we derived macro-structural indicators of sleep quality (e.g., efficiency, duration) and micro-architectural measures of fragmentation and stability (e.g., entropy, sleep bout lengths). To capture autonomic dynamics and activity intensity, we computed a robust set of high-order statistical descriptors, spanning central tendency, variability, and distributional shape (e.g., skewness, kurtosis) for all continuous heart rate, respiratory rate, and activity time series, complemented by specific markers for circadian timing and out-of-bed behavior. Detailed fifty sleep indicators and their explanations are shown in Table S1.

### 2.4 Statistical analysis

We employed a two-part mixed-effects framework to account for the hierarchical structure of the data (nights nested within subjects), the zero-inflated distribution of agitation scores, and systematic cohort differences.

#### 2.4.1. Feature screening and selection

To identify robust physiological predictors while adjusting key covariates, we performed feature screening and selection in two stages. In Stage 1, we assessed the marginal association between each sleep feature (derived from night *j*) and next-day daytime agitation (day *j*+1) using mixed-effects models to account for within-subject correlation. In Stage 2, we addressed redundancy and multicollinearity among candidate predictors via correlation analysis and VIF before fitting the final multivariable mixed-effects models.

##### Agitation Occurrence

We employed a Generalized Linear Mixed Model (GLMM) with a binomial family and logit link function. Let *Y*_*i j*_ denote the binary agitation status (0 or 1) for subject *i* on night *j*, and *p*_*i j*_ = *P*(*Y*_*i j*_ = 1) be the probability of an agitation episode. The model is specified as:

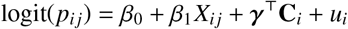

In this equation, *X*_*i j*_ represents the standardized physiological feature of interest (robust z-scored), *β*_0_ denotes the fixed intercept, and *β*_1_ is the regression coefficient quantifying its association with the log-odds of agitation. The term **C**_*i*_ denotes the vector of fixed covariates (standardized age, sex, and data source) with corresponding coefficient vector *γ*, controlling for demographic and device-specific variations. The random intercept 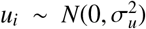 accounts for the subject-specific baseline risk and intra-individual correlation.

##### Agitation Severity

We utilized a Cumulative Link Mixed Model (CLMM) to accommodate the ordinal nature of daily maximum PAS among agitation-positive days (*Y*_*i j*_ ∈ {1, 2, 3, 4}). Unlike linear models that assume equidistant intervals between scores, the CLMM estimates the cumulative probability that the severity falls at or below a specific category *k*:

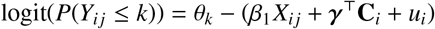

Here, *θ*_*k*_ represents the threshold parameters separating the *k*-th and (*k*+1)-th severity levels, preserving the natural ordering of the PAS scores. The latent physiological effect is subtracted from these thresholds, implying that a positive coefficient *β*_1_ indicates an increased probability of falling into higher severity categories (i.e., exceeding the threshold).

To mitigate the risk of the Type I error rate associated with multiple testing, we applied the Benjamini-Hochberg procedure to control the False Discovery Rate (FDR). Only features meeting a corrected significance threshold of *p* < 0.05 were retained. Subsequently, we addressed multicollinearity among the significant predictors. A Spearman correlation matrix was computed, and for any pair of features with |*r*| > 0.7, the feature with the higher statistical significance (lower *p*-value) was prioritized. A final check using Variance Inflation Factors (VIF) ensured that all retained predictors satisfied VIF < 5.

#### 2.4.2. Multivariable modeling

Building upon the univariate screening phase, we extended the analytical framework to a multivariable context to isolate the independent contributions of the selected physiological markers. This step is critical for disentangling the complex interplay between co-occurring sleep metrics—such as separating the effects of respiratory instability from those of general sleep fragmentation—and determining which markers remain robust predictors after mutual adjustment.

We constructed final multivariable models using the independent features identified in the screening stage. The model specifications mirrored the GLMM and CLMM frameworks defined in Section Feature screening and selection, with the expansion of the single predictor *X*_*i j*_ into a vector of selected independent features **X**_*i j*_. A multivariable GLMM was fitted to the full dataset to estimate adjusted associations with agitation occurrence. A multivariable CLMM was planned for the subset of agitation-positive nights if features survived univariate FDR screening.

#### 2.4.3. Model evaluation

Quantitative results were reported as adjusted Odds Ratios (OR) with 95% Confidence Intervals (CI). To assess the explanatory power of the models, we calculated two goodness-of-fit metrics: Marginal 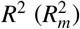, representing the variance explained by the fixed effects (physiological predictors), and Conditional 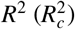, representing the variance explained by the full model including subject-specific random effects. Furthermore, the Intraclass Correlation Coefficient (ICC) was computed to quantify the proportion of variance attributable to between-person (subject-level) differences. Although our primary objective was to estimate adjusted associations, we additionally reported the Area Under the Receiver Operating Characteristic Curve (AUC) as a descriptive summary of the model’s ability to discriminate between agitation and non-agitation nights. Occurrence GLMMs were estimated by variational Bayes in Python 3.13.12 using statsmodels 0.14.6; two-sided normal-approximation p-values were calculated from posterior means and standard deviations. CLMMs were fitted in R 4.5.1 using the ordinal package.

### 2.5 Sensitivity analyses

To assess cross-cohort consistency, we stratified the analysis by device type (WSA vs. EMFIT) and examined whether the direction and magnitude of the pooled associations were similar in the two cohorts.

We also examined sensitivity to the agitation outcome definition. For occurrence, the daily PAS subtype total was defined as the sum of the daily maximum motor, verbal, aggressive, and resistance-to-care scores (range 0–16). Univariate occurrence screening was repeated at daily PAS subtype-total thresholds of ≥2, ≥3, and ≥4. For severity, the univariate CLMM screen was repeated using the daily PAS subtype total as an ordinal outcome among agitation-positive observation pairs.

### 2.6 External validation

To examine whether sleep–agitation associations were also observed in an independent setting, we conducted an external evaluation using the TIHM cohort (*N* = 17). Due to the absence of activity data in this dataset, the evaluation was restricted to the available physiological metrics (heart rate, respiratory rate, and sleep architecture). We fitted adjusted GLMMs controlling for age and sex to estimate associations between the available sleep-derived physiological features and agitation occurrence in this home-living population.

## 3. Results

### 3.1. Descriptive statistics

Table 1 summarizes the study population, data volume, and demographics of the three cohorts. Male patients outnumber female patients across cohorts, and the EMFIT cohort approaches a 1:1 sex ratio. The mean ages for the WSA and EMFIT cohorts are both around 80. Their MMSE (Mini-Mental State Examination) scores are 13.1 (SD = 8.9) and 11.4 (SD = 6.2), respectively, indicating moderate to severe dementia. Due to differences in data collection and annotation protocols, the three cohorts exhibited significant heterogeneity in the proportion of nights with documented next-day agitation.

**Table 1:** Cohort characteristics and night-to-next-day observation pairs.

| Participant-level characteristics |  |  |  |  | Night-to-next-day observation pairs |  |  |
| --- | --- | --- | --- | --- | --- | --- | --- |
| Cohort | Subjects | Age (Mean ± SD) | Diagnosis | MMSE | Agitation-positive pairs | Agitation-negative pairs | Total pairs |
| EMFIT | 21 Male<br>18 Female | 79.7 ± 8.4 | AD: 19<br>Vascular: 5<br>Mixed: 3<br>DLB: 7<br>N/A: 6 | 11.4 ± 6.2<br>(N/A: 5) | 133 | 68 | 201 |
| WSA | 11 Male<br>5 Female | 80.2 ± 8.0 | AD: 10<br>Vascular: 1<br>DLB: 1<br>Other: 1<br>N/A: 3 | 13.1 ± 8.9<br>(N/A: 7) | 101 | 31 | 132 |
| TIHM | 11 Male<br>6 Female<br>(from 56 subjects) | 84.7 ± 8.6 | AD: 40<br>Vascular: 6<br>Other: 9<br>N/A: 1 | N/A | 31 | 770 | 801 |

Note: Night-to-next-day observation pairs link sleep features from night $j$ to the daytime agitation label on day $j + 1$ . Agitation-positive and agitation-negative pairs therefore refer to nights preceding days with and without recorded daytime agitation, respectively. AD = Alzheimer’s disease; DLB = dementia with Lewy bodies; MMSE = Mini-Mental State Examination.

The pooled analytical dataset comprised 333 complete night-to-next-day observation pairs from 55 participants. The daily maximum PAS distribution was 0 in 99 pairs (29.7%), 1 in 91 pairs (27.3%), 2 in 84 pairs (25.2%), 3 in 39 pairs (11.7%), and 4 in 20 pairs (6.0%). The distribution is reported in Supplementary Table S3. Figure 2 summarizes mutually exclusive agitation-domain categories across the 333 night-to-next-day observation pairs. No agitation was recorded in 99 pairs (29.7%). Motor-only agitation occurred in 73 pairs (21.9%), verbal-only agitation in 17 pairs (5.1%), and combined motor and verbal agitation in 139 pairs (41.7%). Other-domain-only agitation, defined as aggression and/or resistance to care without motor or verbal agitation, occurred in 5 pairs (1.5%).

**Figure 2:**
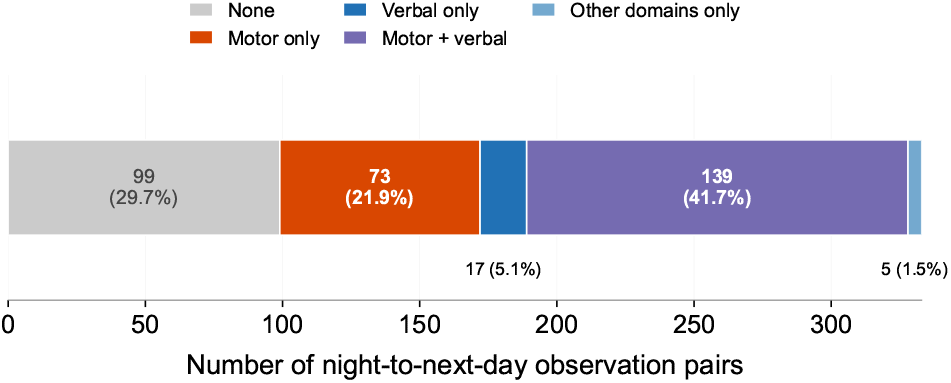
Distribution of mutually exclusive next-day agitation-domain categories across 333 night-to-next-day observation pairs. Categories were defined from daily maximum PAS domain scores. Other-domain-only agitation indicates aggression and/or resistance to care without motor or verbal agitation.

### 3.2. Univariate screening and feature selection for agitation occurrence

#### 3.2.1. All agitation

Initial univariate screening was performed on the pooled samples from the WSA and EMFIT cohorts using GLMM. Six nocturnal features were significant after FDR correction (*p*_*ad j*_ < 0.05); correlation screening retained four nonredundant features for multivariable modeling (Table S2). We observed distinct patterns in respiratory and activity metrics in Figure 3. Lower nocturnal respiratory rates were associated with higher odds of agitation (Min. RR: OR = 0.49, 95% CI: 0.35–0.70; Mean RR: OR = 0.49, 95% CI: 0.32–0.74). In contrast, greater nocturnal activity variability, quantified as the standard deviation of activity, was associated with higher odds of agitation (OR = 1.84, 95% CI: 1.23–2.75). Longer bed presence was also associated with higher odds in univariate screening (OR = 1.65, 95% CI: 1.16–2.34). Supplementary Table S2 reports the complete univariate screening results for all 50 sleep-derived features, including non-significant findings.

**Figure 3:**
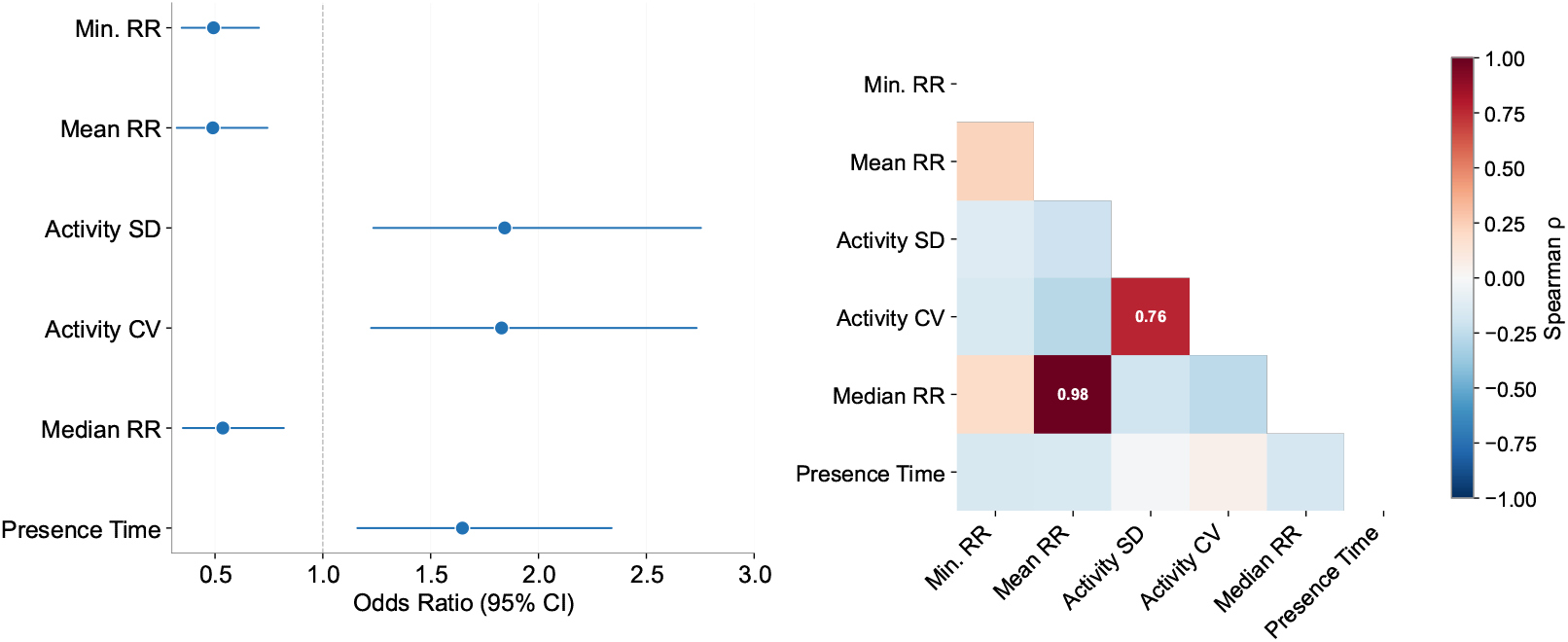
Univariate screening results for agitation occurrence. Effect-size estimates from univariate GLMMs highlight FDR-significant nocturnal features, and the accompanying Spearman correlation heatmap summarizes redundancy among these predictors.

Given the physiological interdependencies among these features, we assessed collinearity before multivariable modeling. Spearman correlation analysis (Figure 3) showed strong redundancy between median RR and mean RR (*r* = 0.98, *p* < 0.001); mean RR was retained because it showed stronger statistical evidence. Similarly, activity SD was prioritized over activity CV (*r* = 0.76). The final feature set comprised minimum and mean RR, activity SD, and presence time. Variance inflation factor (VIF) analysis (Table S2) confirmed negligible multicollinearity among retained predictors (all VIFs ≤ 1.10).

#### 3.2.2. Motor and verbal agitation

Beyond the binary classification of agitation occurrence, we conducted a finer-grained analysis to examine whether agitation subtypes exhibited distinct sleep patterns. Although PAS recorded four subtypes (motor, verbal, resistance to care, and aggression), our analysis focused exclusively on motor and verbal agitation. The prevalence of resistance to care and aggression in our dataset was insufficient to support robust statistical modeling.

Exploratory subtype-specific analyses identified five FDR-significant features for motor agitation (Figure 4). Lower Min. RR (OR = 0.48), minimum HR (OR = 0.56), and minimum activity (OR = 0.50) were associated with lower odds of motor agitation, whereas Activity SD (OR = 1.88) and Activity CV (OR = 1.76) were associated with higher odds. Activity CV was removed from the nonredundant feature set because of its correlation with Activity SD. For verbal agitation, only minimum HR remained significant after FDR correction (OR = 0.47).

**Figure 4:**
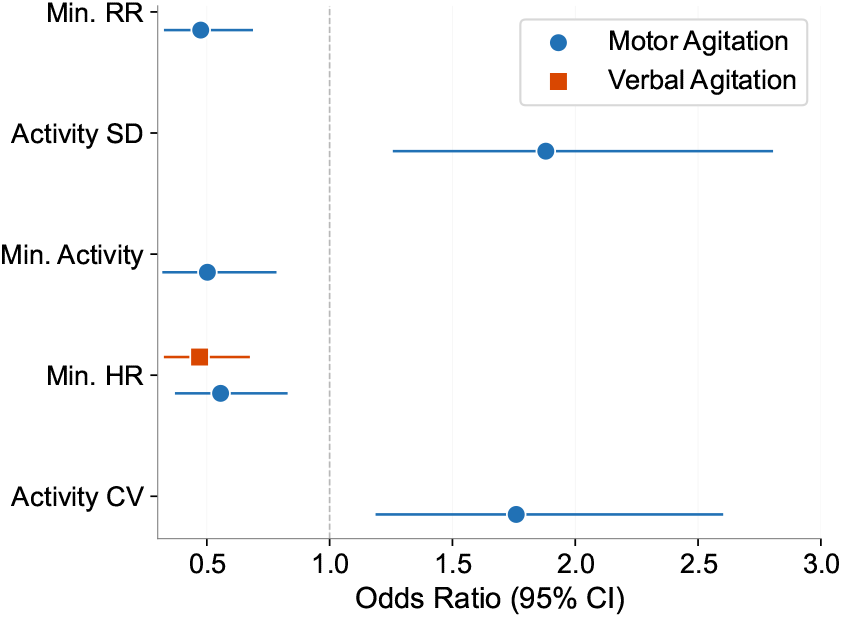
Exploratory subtype-specific analyses. Odds ratios (95% CIs) are shown for features that were FDR-significant in the separate motor- and verbal-agitation GLMMs.

### 3.3. Multivariable predictors of agitation occurrence

After screening individual predictors with univariate GLMM, a multivariable GLMM was fitted to estimate the independent associations of selected sleep features with next-day agitation while accounting for random effects and covariates. As shown in Table 2, two nocturnal features remained significantly associated with the odds of next-day agitation after adjustment for age, sex, and cohort effects.

**Table 2:** Multivariable mixed-effects model for next-day agitation occurrence. Adjusted GLMM estimates (OR, 95% CI) are reported alongside model summary metrics.

| Physiological Features | OR (95% CI) | P-value |
| --- | --- | --- |
| Min. RR ** | 0.58 (0.41 – 0.84) | **0.004** |
| Activity SD * | 1.54 (1.02 – 2.31) | **0.038** |
| Mean RR | 0.69 (0.45 – 1.05) | 0.084 |
| Presence Time | 1.40 (0.97 – 2.01) | 0.071 |
| **Covariates** |  |  |
| Age (per SD) ** | 0.63 (0.45 – 0.89) | **0.009** |
| Sex (Male vs. Female) | 0.99 (0.67 – 1.48) | 0.976 |
| Cohort (WSA vs. EMFIT) *** | 2.68 (1.59 – 4.51) | < 0.001 |
| **Model Performance** |  |  |
| Samples / Subjects | 333 / 55 |  |
| ICC | 0.360 |  |
| Marginal $R^2$ | 0.178 | |
| Conditional $R^2$ | 0.474 | |
| AUC | 0.707 |  |

Note: AUC = Area Under the ROC Curve; CI = Confidence Interval; ICC = Intraclass Correlation Coefficient; Min. = Minimum; OR = Odds Ratio; RR = Respiratory Rate; SD = Standard Deviation. ORs are adjusted for age, sex, and cohort. \* indicates statistical significance ( $p < 0.05$ ). Reference categories are Female for Sex and EMFIT for Cohort.

In the multivariable model, higher Min. RR was independently associated with lower odds of next-day agitation (OR = 0.58, 95% CI: 0.41–0.84, *p* = 0.004), whereas greater Activity SD was associated with higher odds (OR = 1.54, 95% CI: 1.02–2.31, *p* = 0.038). Mean RR (OR = 0.69, *p* = 0.084) and presence time (OR = 1.40, *p* = 0.071) did not retain statistical significance after mutual adjustment.

Regarding covariates, WSA observations had higher adjusted odds of agitation than EMFIT observations (OR = 2.68, *p* < 0.001). Higher age was associated with lower odds per SD increase (OR = 0.63, *p* = 0.009), while sex was not significant (*p* = 0.976).

The multivariable model had an in-sample AUC of 0.707. Fixed effects explained 17.8% of the variance in agitation occurrence 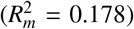, while the full model explained 47.4% 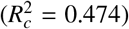. The gap between these estimates, together with an ICC of 0.360, indicates substantial between-subject heterogeneity in agitation likelihood.

### 3.4. Exploratory univariate screening for agitation severity

In the second stage of the two-part model, we analyzed agitation intensity within the subset of nights preceding next-day agitation events (*N* = 234 nights from 52 patients).

Seven sleep features had nominal *p* < 0.05 in the univariate CLMM analysis, but none remained significant after FDR correction (smallest adjusted *p* = 0.096; Figure 5). Because no feature survived the prespecified screening step, no feature-based multivariable CLMM was fitted. A separate covariate-only baseline CLMM was retained to summarize demographic, cohort, and subject-level variation (Table 3).

**Table 3:**
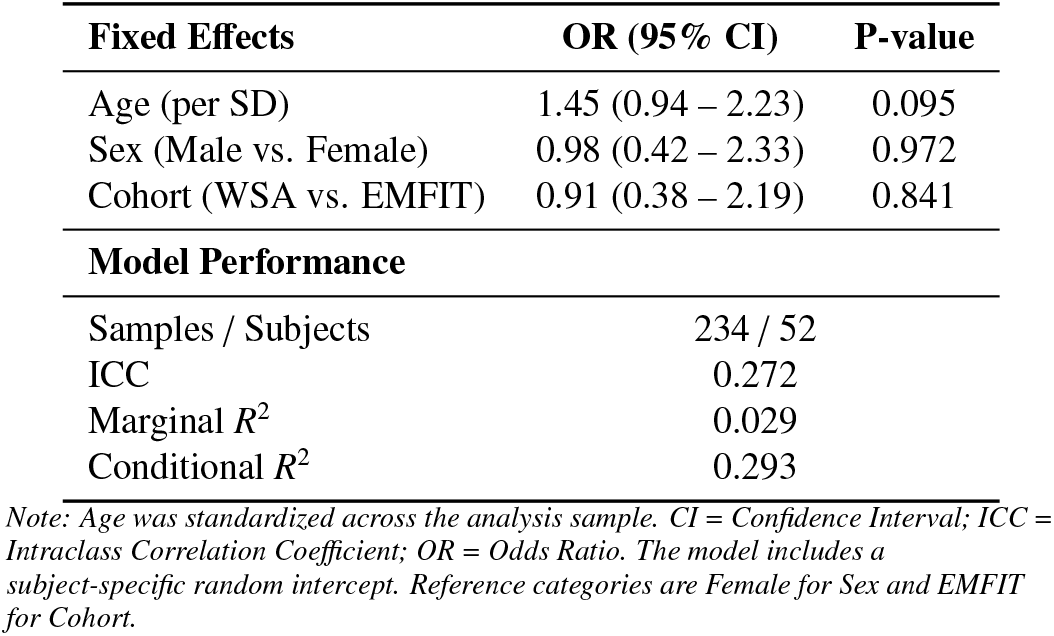
Baseline severity model among agitation-positive days. Covariate-only CLMM estimates summarize the association of demographic and cohort factors with severity.

| Fixed Effects | OR (95% CI) | P-value |
| --- | --- | --- |
| Age (per SD) | 1.45 (0.94 – 2.23) | 0.095 |
| Sex (Male vs. Female) | 0.98 (0.42 – 2.33) | 0.972 |
| Cohort (WSA vs. EMFIT) | 0.91 (0.38 – 2.19) | 0.841 |
| **Model Performance** |  |  |
| Samples / Subjects | 234 / 52 |  |
| ICC | 0.272 |  |
| Marginal $R^2$ | 0.029 | |
| Conditional $R^2$ | 0.293 | |

Note: Age was standardized across the analysis sample. CI = Confidence Interval; ICC = Intraclass Correlation Coefficient; OR = Odds Ratio. The model includes a subject-specific random intercept. Reference categories are Female for Sex and EMFIT for Cohort.

**Figure 5:**
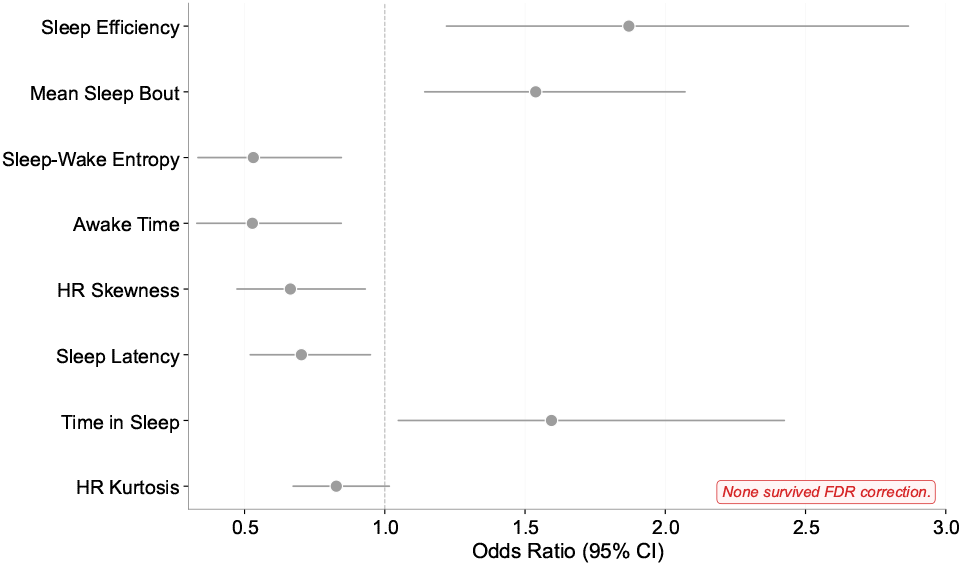
Association between sleep features and agitation severity. Across agitation-positive days, no sleep feature showed a robust association with severity after FDR correction.

None of the baseline covariates was significantly associated with agitation severity. Fixed effects explained 2.9% of the variAnce 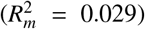, while the full baseline model explained 29.3% 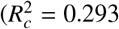; ICC = 0.272).

### 3.5. Sensitivity analysis

#### 3.5.1. WSA-only and EMFIT-only GLMMs

We fitted cohort-specific univariate GLMMs for WSA and EMFIT (Figure S1). No feature remained significant after FDR correction in either cohort. The pooled respiratory-rate associations had ORs < 1 in both cohort-specific analyses, while Activity SD, Activity CV, and presence time had ORs > 1 in both cohorts.

#### 3.5.2. Sensitivity to agitation outcome definitions

Outcome-definition sensitivity results are summarized in Supplementary Table S3. At daily PAS subtype-total thresholds of ≥2, ≥3, and ≥4, 181 (54.4%), 135 (40.5%), and 86 (25.8%) of the 333 observation pairs were positive, respectively. The retained features were sleep latency at ≥2, minimum HR and mean wake-bout length at ≥3, and OOB count at ≥4. For agitation severity, neither daily maximum PAS nor daily PAS subtype total yielded an FDR-significant sleep feature in the univariate CLMM screens.

### 3.6. External validation on TIHM cohort

Univariate GLMM results for TIHM are shown in Figure S2. Higher mean RR was associated with lower odds of agitation (OR = 0.44, 95% CI: 0.25–0.76, FDR-adjusted *p* = 0.045). RR SD (OR = 0.49, 95% CI: 0.36–0.67, FDR-adjusted *p* < 0.001) and RR CV (OR = 0.51, 95% CI: 0.34–0.77, FDR-adjusted *p* = 0.029) were also significant, with feature-level AUCs ranging from 0.64 to 0.69. These results provide external support for respiration-related associations with agitation occurrence.

## 4. Discussion

### 4.1. Principle findings

This study utilized contactless under-mattress sensors to examine associations between nocturnal features and next-day agitation in real-world dementia settings. Six features remained significant after FDR correction: minimum RR, mean RR, median RR, Activity SD, Activity CV, and presence time. After correlation screening, the four nonredundant features retained for multivariable modeling were minimum RR, mean RR, Activity SD, and presence time. In the multivariable analysis, minimum RR and Activity SD were independently associated with agitation occurrence. These observational findings identify sleep-derived correlates of next-day agitation occurrence and motivate further investigation of the physiological pathways linking nocturnal sleep and daytime behavior.

#### The physiological signature of agitation risk

The independent associations of Min. RR and Activity SD suggest that night-to-night fluctuations in physiological regulation are associated with next-day agitation risk. Specifically, a lower nocturnal respiratory baseline was independently associated with a higher likelihood of next-day agitation. Given the tight coupling between respiratory rhythms and autonomic regulation [31, 32], this pattern may reflect altered autonomic tone, which has been linked to agitation and broader behavioral dysregulation in AD and related neurodegenerative conditions [33]. Furthermore, this interpretation aligns with recent in-home monitoring work in which respiration-related features contributed to identifying agitation episodes [24].

Elevated nocturnal activity instability may indicate increased nighttime restlessness and poorer sleep continuity. This interpretation is consistent with prior literature in which movement-derived indices are used as behavioral proxies of fragmented sleep [34, 35], and echoes earlier evidence linking disrupted sleep continuity to daytime agitation in PLWD [36].

More broadly, our findings fit within established models of sleep-dependent emotional regulation, which posit that sleep disruption can reduce prefrontal regulatory influence and heighten next-day emotional reactivity [13]. Moreover, disrupted sleep continuity has been associated with diminished emotional and executive regulation in dementia populations [37], linking sleep disruption to adverse emotional, behavioral, and cognitive outcomes.

Notably, although prolonged time in bed (Presence Time) was significant in univariate analysis, it did not remain significant in the multivariable model. While extended time in bed is a common feature of disturbed sleep in people living with dementia [23], it is a relatively coarse indicator that may also reflect broader factors such as reduced mobility, functional impairment, or care routines, rather than the proximal nocturnal processes most relevant to agitation risk. In contrast, our results suggest that next-day agitation is more closely associated with markers of disrupted sleep continuity and nocturnal physiological dysregulation during the time spent in bed.

#### Differential sleep–agitation associations across phenotypes and outcomes

Exploratory subtype-specific analyses identified five FDR-significant features for motor agitation and one for verbal agitation. This pattern is consistent with Cohen-Mansfield’s “Unmet Needs Model”, which proposes that behavioral symptoms may arise from distinct underlying etiologies [7]. Motor agitation may be more closely related to nocturnal restoration and physiological instability, whereas verbal agitation may also reflect proximal psychosocial factors such as distress, pain, unmet needs, or environmental deprivation. Similar subtype-dependent physiological patterns and patient-level heterogeneity have been reported in wearable-sensing studies of agitation [38].

Furthermore, the two-part analysis showed a differential pattern between agitation occurrence and intensity. No sleep feature remained associated with severity after FDR correction, so no feature-based multivariable severity model was fitted. In the covariate-only baseline model, fixed effects explained little variance, while the ICC indicated appreciable subject-level heterogeneity. This pattern is consistent with the possibility that episode severity depends more on factors such as disease stage, temperament, or premorbid personality [39, 40].

The occurrence sensitivity analyses identified different retained features across daily PAS subtype-total thresholds, indicating that the associated feature profile varies with the definition of accumulated agitation burden. For severity, no FDR-significant sleep feature was identified using either daily maximum PAS or daily PAS subtype total. Thus, the occurrence results varied across agitation burden thresholds, while the FDR-level severity finding was consistent across the two aggregation definitions.

### 4.2. Clinical implications and robustness

The observed night-to-next-day associations support prospective evaluation of passive sleep sensing for short-term risk stratification. Key next steps are to determine whether sleep-derived markers add information beyond usual clinical observation and whether that information can support timely care decisions.

We adopted a two-part framework, using GLMM for occurrence and CLMM for severity, to separate these two outcomes. The pooled associations had consistent directions in the WSA and EMFIT analyses, and respiration-related associations were also observed in the independent TIHM home-monitoring cohort. The lower agitation prevalence in TIHM is consistent with its community setting, exclusion criteria, sparse event-alert labeling, and binary validation workflow. The TIHM analysis extends evaluation of respiration-related associations to a different setting and label mechanism.

### 4.3. Limitations and future work

Several limitations merit consideration. First, the potential “sedative effect” of psychotropic medications may confound the relationship between sleep efficiency and agitation severity, as sedatives can influence sleep metrics while masking or exacerbating behavioral symptoms through disinhibition. Future analyses must account for pharmacological treatments to disentangle these effects. Moreover, while our dense labeling strategy improves upon spot-checks and we employed daily aggregation to mitigate label noise, the behavioral ground truth remains subject to observer bias.

The primary cohort included several dementia diagnoses. Accordingly, the reported estimates describe population-level associations across this heterogeneous cohort rather than diagnosis-specific effects. The small and unevenly represented diagnostic subgroups did not support reliable diagnosis-stratified estimation [41]. Future studies with larger diagnosis-specific samples can evaluate whether sleep–agitation associations differ by dementia subtype. Longer within-person monitoring can assess the temporal stability of night-to-next-day associations and characterize person-specific patterns. Prospective studies can also evaluate whether sleep-derived risk information adds value beyond routine observation for care decisions and patient outcomes.

Future research should focus on deploying multimodal monitoring systems that combine nocturnal physiology with objective daytime activity tracking (e.g., wearables) and environmental sensing (noise, light) to capture the full 24-hour behavioral cycle. Furthermore, integrating medication timing data is essential to disentangle physiological sleep regulation from drug-induced sedation. Validating these biomarkers in larger, medication-controlled trials is a critical step toward translating these findings into real-time, personalized early warning systems for dementia care.

### 4.4 Conclusion

This study provides evidence that contactless nocturnal sleep features, particularly respiratory-rate and activity-variability markers, are associated with next-day agitation occurrence in dementia care settings. In the multivariable analysis, lower minimum RR and greater Activity SD were independently associated with agitation occurrence, whereas no sleep feature remained associated with severity after FDR correction. These findings provide a basis for prospective evaluation of sleep-informed short-term risk stratification in dementia care.

## Acknowledgments

The authors thank all participating patients and their families, as well as the nursing staff and psychiatrists in training at Cog K of UPC KU Leuven.

## 6. Author contributions

**Zhen Liu:** Conceptualization, Methodology, Software, Investigation, Data Curation, Writing - original draft. **Marta Bono:** Investigation, Data Curation, Writing - review & editing. **Ajda Flisar:** Investigation, Data Curation, Writing - review & editing. **Robbe Decloedt:** Investigation, Data Curation, Writing - review & editing. **Maarten Van Den Bossche:** Conceptualization, Supervision, Writing - review & editing. **Maarten De Vos:** Conceptualization, Supervision, Writing - review & editing.

## 7. Statements and declarations

### 7.2. Consent to participate

Informed consent was obtained from all individual participants included in the study. For participants who were unable to provide consent due to cognitive impairment, written informed consent was obtained from their legal representatives or primary caregivers. All procedures were performed in accordance with the study protocol approved by the Research Ethics Committee of University Hospitals Leuven.

### 7.3. Consent for publication

Informed consent for the publication of de-identified data and study results was obtained from all participants (or their legal representatives/caregivers) as part of the recruitment and consent process.

### 7.4. Declaration of conflicting interest

The authors declare that they have no known competing financial interests or personal relationships that could have appeared to influence the work reported in this paper.

### 7.5. Funding statement

This work was supported by the Flanders AI Research (FAIR) Program; the Research Foundation – Flanders (FWO) under grant number G046925N; the ‘Funds Malou Malou, Perano, Georgette Paulus, JMJS Breugelmans and Gabrielle, François and Christian De Mesmaeker’ managed by the King Baudouin Foundation of Belgium (2021-J1990130-222081); KU Leuven (grants C2M/23/053, CELSA/24/019, and the Global PhD partnerships with the University of Melbourne GPUM/22/022); the Paris Brain Institute and KU Leuven (Big Brain Theory 4 grant); the ‘Klinische onderzoeksen opleid-ingsraad (KOOR)’ of the University Hospitals Leuven; and the AI-PROGNOSIS project under the European Union’s Horizon Europe research and innovation program (Grant Agreement No. 101080581). Views and opinions expressed are however those of the author(s) only and do not necessarily reflect those of the European Union. Neither the European Union nor the granting authority can be held responsible for them.

### 7.6. Data availability

The primary dataset analyzed in the current study is not publicly available due to patient privacy and ethical restrictions, but de-identified data are available from the corresponding author upon reasonable request. The secondary TIHM dataset used in this study is publicly available on Zenodo at: https://zenodo.org/records/7622128.

### 7.7. Declaration of generative AI use

During the preparation of this work, the authors used Google’s Gemini for English editing, code debugging, and optimization during manuscript preparation. All content was reviewed and verified by the authors, who retain full responsibility for the final work.

## Supplementary Material

**Table S1:** Night-level features extracted from time-series sensor data. Summary of candidate sleep-derived features (respiratory, cardiac, movement, and sleep-timing proxies) computed per night for downstream screening and modeling.

| Feature | Description |
| --- | --- |
| Time in bed | Total time in bed (minutes) |
| Time in sleep | Total sleep duration (minutes) |
| Presence time | Total duration between the first bed entry and the final bed exit (minutes) |
| Sleep efficiency | Time in sleep / Time in bed |
| Awake time | Awake time during presence (minutes) |
| WASO | Wake after sleep onset (minutes) |
| Awake ratio | WASO / total sleep time |
| Awakenings | Number of awakenings after first sleep onset |
| Sleep latency | Time from on-bed to first sleep (minutes) |
| Mean sleep bout length | Mean continuous sleep bout length (minutes) |
| Mean wake bout length | Mean continuous wake bout length (minutes) |
| Sleep-wake entropy | Shannon entropy of sleep-wake state transitions |
| HR-RR correlation in sleep | Spearman correlation between HR and RR during sleep |
| Sleep midpoint (sin) | Sine component of midsleep time (circular encoding) |
| Sleep midpoint (cos) | Cosine component of midsleep time (circular encoding) |
| OOB count | Number of Out-of-Bed (OOB) events during sleep period |
| OOB duration | Total duration of OOB events during sleep period (minutes) |
| OOB percent | OOB percentage (OOB duration/presence duration) |
| Average OOB time | Average duration per OOB event (minutes) |
| Long OOB Flag | Flag indicating if max OOB duration $\geq 20$ minutes (1=yes, 0=no) |
| Statistics of HR, RR, Activity | Mean, Median, Min, Max, SD, CV, IQR, MAD, Skewness, Kurtosis |

**Table S2:**
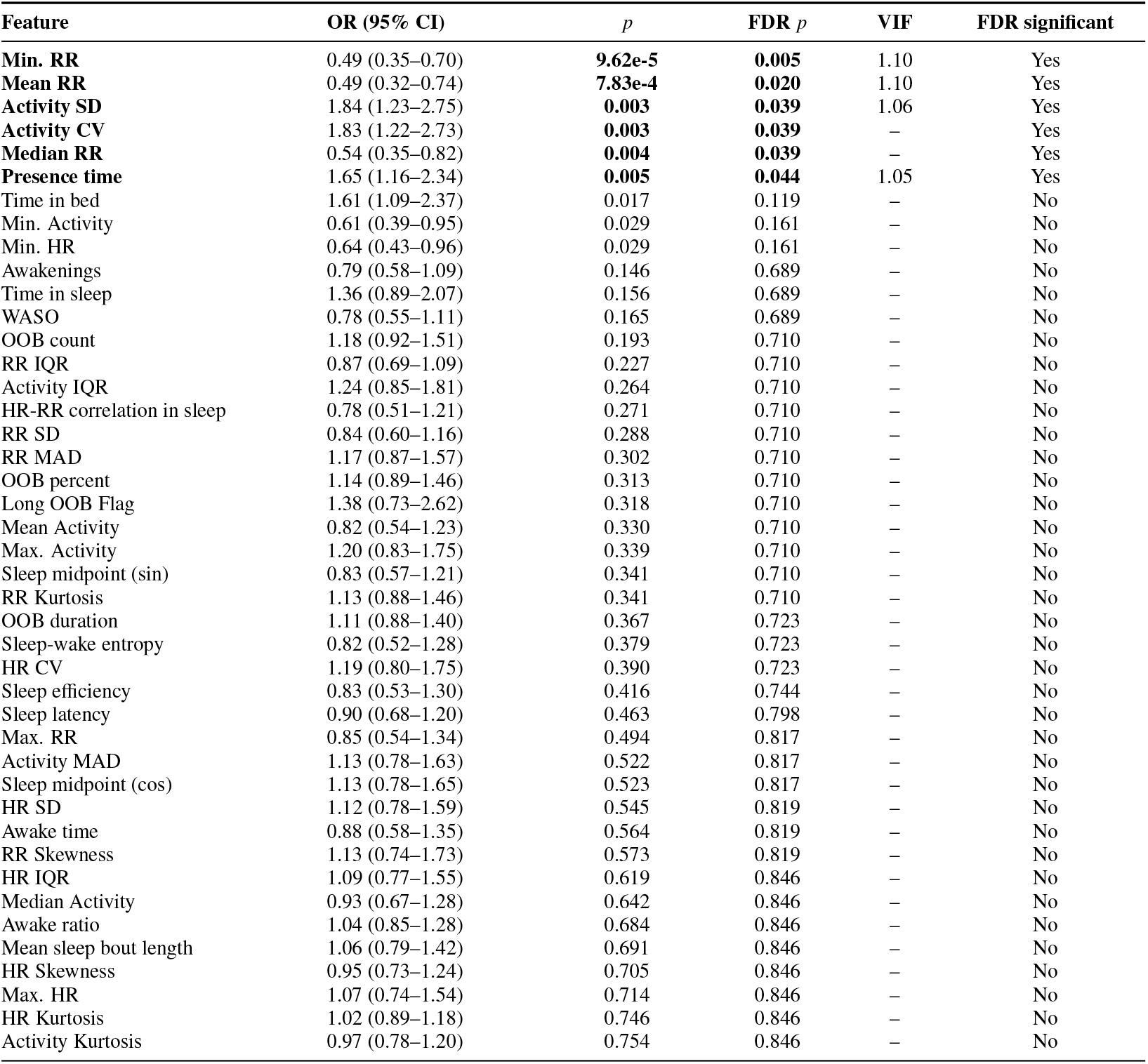
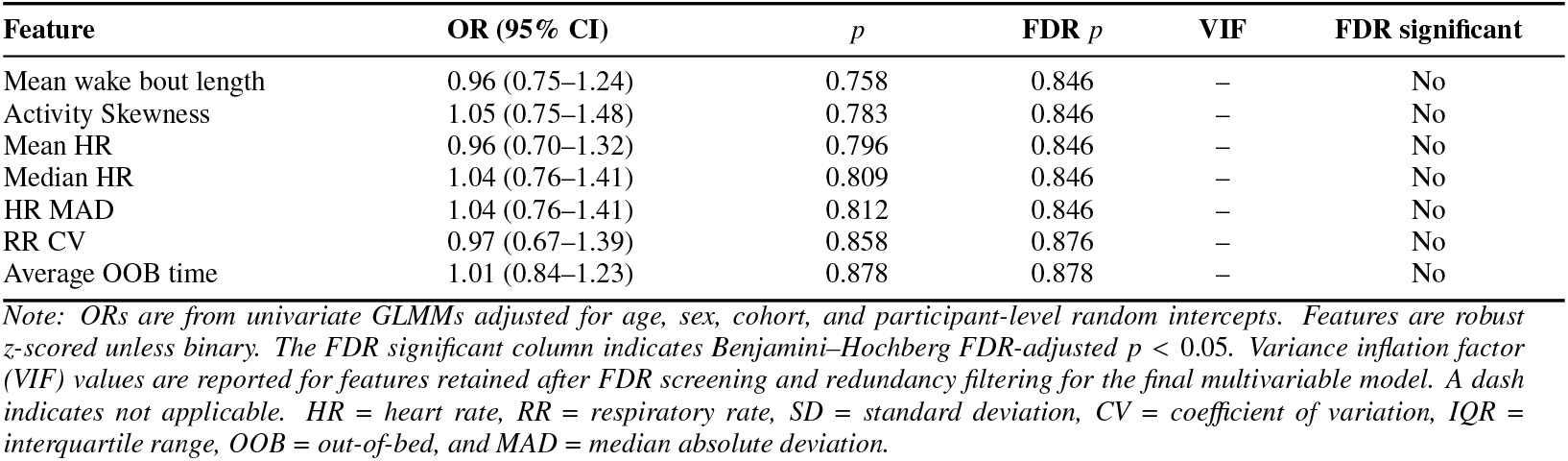
Complete univariate GLMM screening for next-day agitation occurrence across all 50 sleep-derived features.

**Table S3:** PAS distribution and outcome-definition sensitivity analyses for agitation occurrence and severity.

| Daily maximum PAS distribution across 333 night-to-next-day observation pairs |  |  |
| --- | --- | --- |
| Daily maximum PAS | Observation pairs | Percentage |
| 0 | 99 | 29.7% |
| 1 | 91 | 27.3% |
| 2 | 84 | 25.2% |
| 3 | 39 | 11.7% |
| 4 | 20 | 6.0% |
| Occurrence-definition sensitivity analyses |  |  |
| Occurrence definition | Positive pairs (%) | FDR-significant and retained features |
| Any PAS subtype > 0 | 234 (70.3%) | Six FDR-significant features. Retained: Min. RR (OR = 0.49, 95% CI: 0.35–0.70), Mean RR (OR = 0.49, 95% CI: 0.32–0.74), Activity SD (OR = 1.84, 95% CI: 1.23–2.75), and Presence time (OR = 1.65, 95% CI: 1.16–2.34). Complete results are reported in Supplementary Table S2. |
| Daily PAS subtype total ≥ 2 | 181 (54.4%) | One FDR-significant feature. Retained: Sleep latency (OR = 0.58, 95% CI: 0.43–0.78). |
| Daily PAS subtype total ≥ 3 | 135 (40.5%) | Two FDR-significant features. Retained: Min. HR (OR = 0.43, 95% CI: 0.29–0.64) and mean wake-bout length (OR = 0.63, 95% CI: 0.49–0.81). |
| Daily PAS subtype total ≥ 4 | 86 (25.8%) | Three FDR-significant features. Retained: OOB count (OR = 1.40, 95% CI: 1.18–1.66). |
| Severity sensitivity analyses |  |  |
| Severity outcome | Analysis | FDR-significant sleep features |
| Daily maximum PAS | CLMM screen | None |
| Daily PAS subtype-total | CLMM screen | None |

Note: Daily maximum PAS is the highest of the daily maximum motor, verbal, aggressive, and resistance-to-care scores (range 0–4). Daily PAS subtype total is the sum of these four subtype maxima (range 0–16). Occurrence sensitivity analyses applied thresholds to the daily PAS subtype total. The severity sensitivity analysis treated the same total as an ordinal outcome among agitation-positive observation pairs. OOB = out-of-bed.

**Figure S1:**
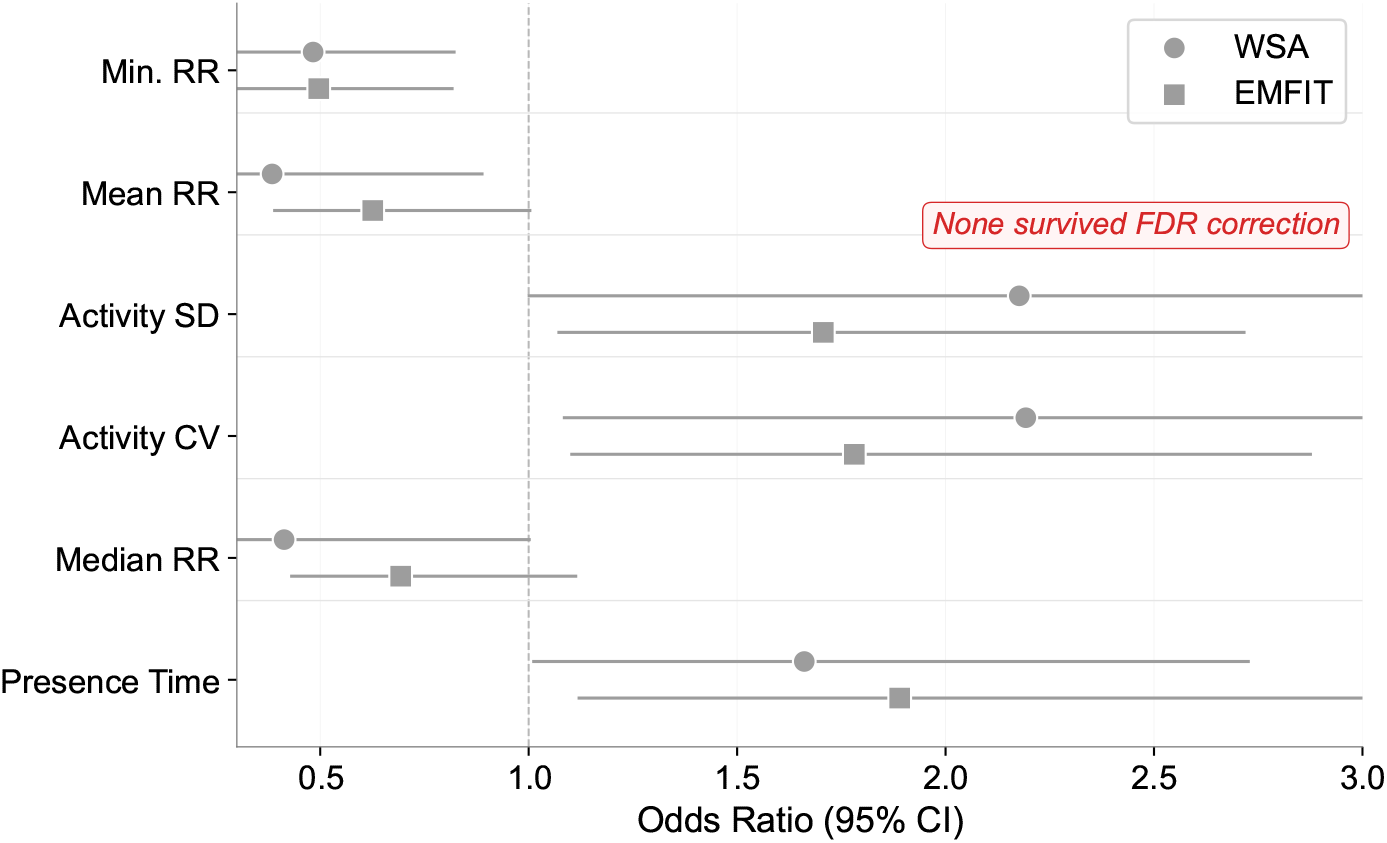
Device-specific sensitivity analysis of agitation occurrence. Adjusted odds ratios (95% CIs) from separate univariate GLMMs in WSA and EMFIT are shown for the six features that were FDR-significant in pooled screening. Effect directions were consistent across cohorts; no feature was FDR-significant in either cohort-specific analysis.

**Figure S2:**
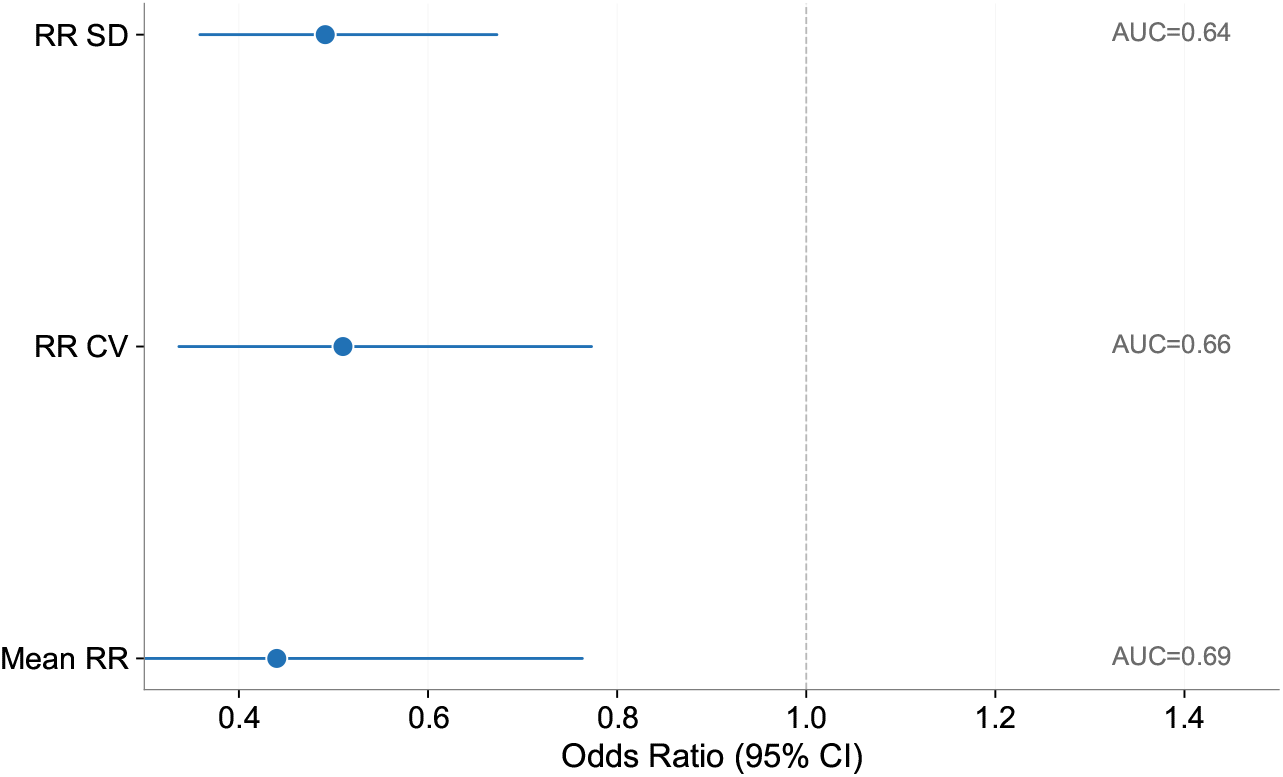
External evaluation in TIHM. FDR-significant respiration-related associations from adjusted single-feature GLMMs are shown with odds ratios, 95% CIs, and feature-level AUCs. CV = Coefficient of Variation; SD = Standard Deviation.

## Notes

### Competing Interest Statement

The authors have declared no competing interest.

### Author Declarations

The Ethics Committee Research of University Hospitals Leuven gave ethical approval for this work (ID: S62882).

### Summary of Updates

The title and selected wording were revised for precision, and additional details on the data, outcome definitions, and sensitivity analyses were added.

